# Decline in prenatal buprenorphine/naloxone fills during the COVID-19 pandemic in the United States

**DOI:** 10.1101/2021.10.08.21264760

**Authors:** Ashley L. O’Donoghue, Alyse Reichheld, Timothy S. Anderson, Chloe A. Zera, Tenzin Dechen, Jennifer P. Stevens

**Author notes:** **Corresponding author:** Ashley L. O’Donoghue, PhD, Center for Healthcare Delivery Science, Beth Israel Deaconess Medical Center, 330 Brookline Avenue, Boston, MA 02215.

## Abstract

**Background and Aims:** Pregnancy provides a critical opportunity to engage women with substance use disorder in care. Buprenorphine/naloxone treatment is associated with improved pregnancy and fetal outcomes, but prior to the COVID-19 pandemic, there were multiple barriers to accessing buprenorphine/naloxone during pregnancy. Care disruptions during the pandemic may have further exacerbated these already existing barriers. To quantify these changes, we examined trends in the number of individuals filling prescriptions for prenatal buprenorphine/naloxone prescriptions during the COVID-19 pandemic.

**Methods:** We estimated an interrupted time series model using linked national pharmacy claims and medical claims data from May 2019 to December 2020. We estimated changes in the level and trend in the monthly number of individuals filling prescriptions for prenatal buprenorphine/naloxone during the COVID-19 pandemic. We then stratified our analyses by payer.

**Results:** We identified 2,947 pregnant patients filling buprenorphine/naloxone prescriptions. Before the pandemic, there was positive growth in the monthly number of individuals filling prescriptions for prenatal buprenorphine/naloxone (4.83% (95% confidence interval (CI): 3.40% to 6.26%). During the pandemic, the monthly growth rate in individuals filling prescriptions for prenatal buprenorphine/naloxone declined for both patients on commercial insurance and patients on Medicaid (all payers: -5.53% (95% CI: -7.28% to -3.78%); Medicaid: -7.66% (95% CI: -10.42% to -4.90%); Commercial: -3.59% (95% CI: -5.53% to -1.66%)).

**Conclusion:** The number of pregnant individuals filling buprenorphine/naloxone prescriptions was increasing prior to the pandemic, but this growth has been lost during the pandemic.

## INTRODUCTION

The number of pregnant patients with opioid use disorder (OUD) has increased four-fold in the past decade^1^. The prenatal use of medication for opioid use disorder, such as buprenorphine/naloxone, is associated with improved prenatal care adherence and pregnancy outcomes, including lower rates of preterm birth and low birth weight, as well as decreased rates of maternal relapse and overdose^2,3,4^. Despite this, before the COVID-19 pandemic, only 55% of pregnant patients with OUD were on medication for their opioid use^5^.

Prior to the COVID-19 pandemic, there were multiple barriers to prenatal buprenorphine/naloxone access, including limited access to providers with obstetric and addiction treatment expertise, stigma, and fear of legal or child welfare consequences ^6,7^. To circumvent potential loss of access to care during the COVID19 pandemic, the federal government permitted the initiation of buprenorphine/naloxone treatment via telehealth in June 2020.

Pregnancy offers a critical opportunity to engage women with OUD in care, but it is unclear how the COVID-19 pandemic affected access to buprenorphine/naloxone in this population. We use an interrupted time series (ITS) design to examine trends in the number of individuals filling prescriptions for prenatal buprenorphine/naloxone during the COVID-19 pandemic.

## METHODS

We used Symphony Health’s pharmacy claims and medical claims database, which covers over 200 million patients^8^. We analyzed monthly trends in the number of pregnant patients filling buprenorphine/naloxone prescriptions from May 2019 to December 2020. We defined prenatal buprenorphine/naloxone fills as prescription fills in the six months prior to delivery. Specifically, we extracted all delivery visits from Symphony Health’s medical claims database using CPT codes 59409, 59612, 59514, and 59620. These patients were then linked to prescription claims data, and patients with at least one buprenorphine/naloxone fill in the six months prior to delivery were included.

We were concerned that there might be seasonality or changes in pregnancy trends during the pandemic. To adjust for this potential variation, we calculated the total number of currently pregnant patients in each month as the number of patients with a delivery date in the subsequent six months. We then weighted the number of prenatal buprenorphine/naloxone fills by the total number of pregnancies each month. We followed the same process for the analyses stratified by payer to account for differences in pregnancy trends by payer. That is, we calculated the total number of currently pregnant patients each month on Medicaid and the total number of currently pregnant patients each month on commercial insurance. Then, we weighted the monthly count of prenatal buprenorphine/naloxone fills for Medicaid patients and the monthly count of prenatal buprenorphine/naloxone fills for commercially insured patients by the total monthly number of pregnancies for that payer.

We compared the pre-pandemic (May 2019 to February 2020) level and growth rate to the post-pandemic (April 2020 to December 2020) level and growth rate of the number of weighted individuals filling prescriptions for prenatal buprenorphine/naloxone. Predicted levels are calculated from a linear regression of monthly trends on the weighted number of individuals filling prescriptions for prenatal buprenorphine/naloxone. Predicted growth rates are calculated from a log-linear regression of monthly trends on the log-transformed weighted number of individuals filling prescriptions for prenatal buprenorphine/naloxone. We estimated the change in level and growth rate using an interrupted time series (ITS) design. The change in level from pre- to post-pandemic is calculated as the coefficient on the term for the breakpoint in the ITS regression. The change in trend is calculated as the coefficient on the interaction term in the ITS regression where the dependent variable is log-transformed. We stratified the ITS analysis by payer.

We used Stata SE 16 (StataCorp) for statistical analysis, and a p-value < 0.05 was considered statistically significant. This research was classified as exempt by the Beth Israel Deaconess Medical Center institutional review board.

## RESULTS

We identified 2,947 pregnant patients filling buprenorphine/naloxone prescriptions. Most patients (55.5%) were between the ages of 21 and 30 and most patients (58.1%) were on commercial insurance while 38.9% were on Medicaid.

The Figure displays monthly trends in the number of individuals filling prescriptions for prenatal buprenorphine/naloxone per 100,000 pregnancies by payer. Prior to the pandemic, the monthly growth rate in the number of individuals filling prescriptions for prenatal buprenorphine/naloxone was positive overall and across payers (all payers: 4.83% (95% confidence interval (CI): 3.40% to 6.26%); Medicaid: 5.35% (95% CI: 3.44% to 7.25%); Commercial: 4.06% (95% CI: 2.65% to 5.48%)). There was no immediate statistically significant change in the number of individuals filling prenatal buprenorphine/naloxone at the beginning of the pandemic. However, as the pandemic continued, the monthly growth rate declined by 5.53% (95% CI: -7.28% to -3.78%). There was a decline among both patients on Medicaid (−7.66%, 95% CI: -10.42% to -4.90%), and patients on commercial insurance (−3.59%, 95% CI: -5.53% to -1.66%).

**Figure.**
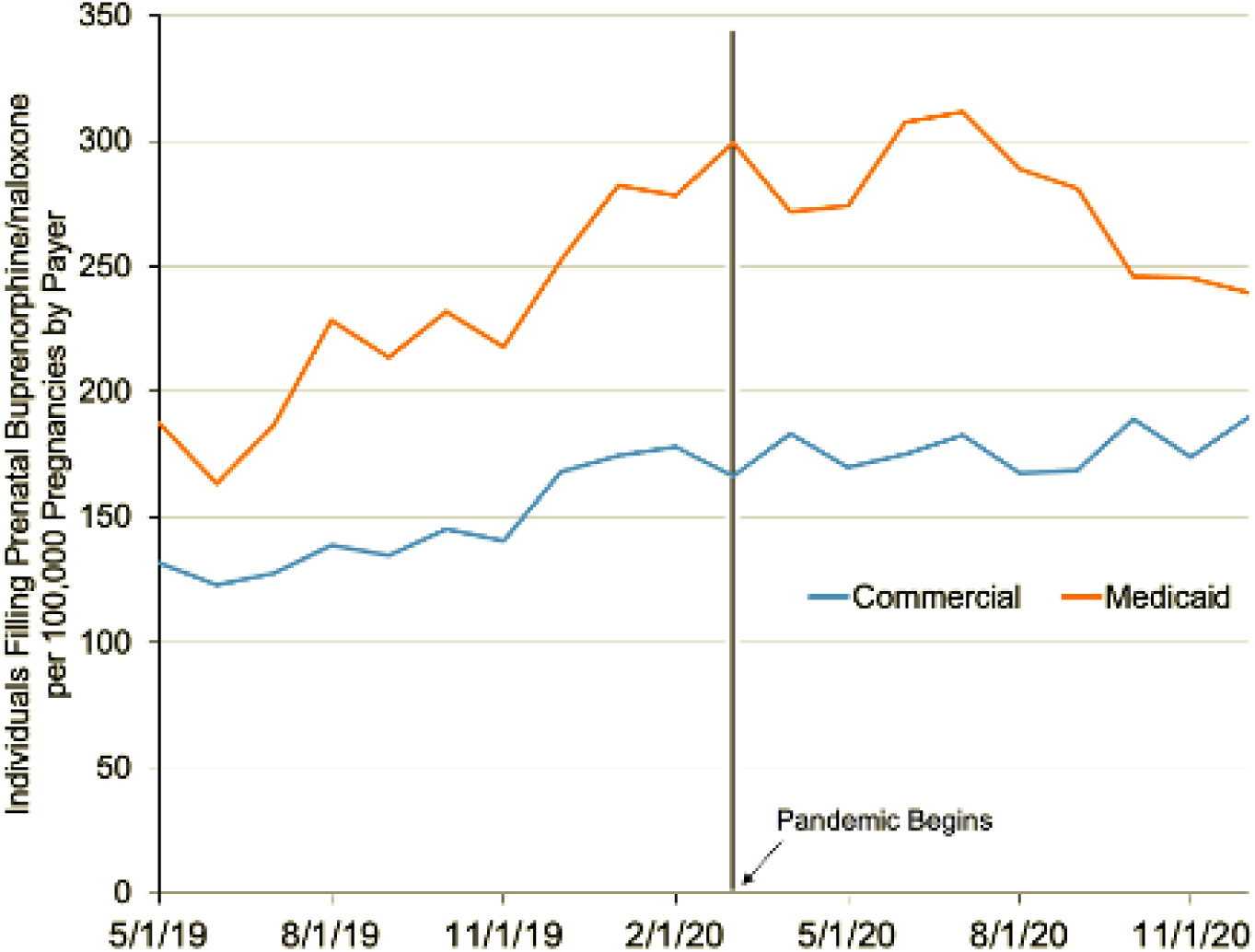
Trends in the number of individuals filling prenatal buprenorphine/naloxone prescriptions by payer. Trends in individuals filling prenatal buprenorphine/naloxone prescriptions each month from Symphony Health claims data from May 2019 to December 2020. The y-axis represents the number of individuals filling prenatal buprenorphine/naloxone prescriptions, weighted by the total number of pregnancies for a given month and payer. This weighting is to account for any seasonality in pregnancies or changing trends in pregnancies during the pandemic that may differ over time and by payer. The vertical black line denotes March 2020, when the pandemic began in the United States.

## DISCUSSION

Our findings suggest that, prior to the COVID-19 pandemic, the number of pregnant individuals filling buprenorphine/naloxone prescriptions was increasing. This growth, however, has been lost during the pandemic, across both patients on commercial insurance and Medicaid.

There are several limitations to this study. First, our sample size of pregnant patients filling prenatal buprenorphine/naloxone prescriptions is relatively small. Second, we are not able to determine gestation age at time of delivery, and so we defined prenatal buprenorphine/naloxone fills as any fill within the six months prior to delivery. As a result, we could be missing patients who filled buprenorphine/naloxone prescriptions early in a full-term pregnancy and then discontinued treatment during the first trimester. Conversely, we could be including women who delivered at less than six months gestation and filled a buprenorphine/naloxone prescription prior to, rather than during, a pregnancy.

Our findings highlight an exacerbated barrier in access to prenatal buprenorphine/naloxone treatment during the COVID-19 pandemic. Future studies could explore whether these declines indicate impaired access to prenatal care, decreased initiation of buprenorphine/naloxone treatment among pregnant patients with OUD, or diminished adherence to treatment during the pandemic. Pregnancy is a critical opportunity to engage women with OUD in treatment, but our findings suggest that we may be missing this chance more often during the pandemic.

## Data Availability

The data, technology, and services used in the generation of these research findings are restricted access and were generously supplied pro bono by the COVID-19 Research Database partners. Applications for access can be found at the following URL: https://covid19researchdatabase.org/.

## ACKNOWLEDGEMENTS

### Funding

O’Donoghue, Dechen, and Stevens are funded by an unrestricted philanthropic gift from Google.org.

## Acknowledgements

The data, technology, and services used in the generation of these research findings were generously supplied pro bono by the COVID-19 Research Database partners, who are acknowledged at https://covid19researchdatabase.org/.

---

**Table:** Change in Monthly Individuals Filling Prenatal Buprenorphine Prescriptions per 100,000 pregnancies by payer^a^ during the COVID-19 Pandemic^b^.

|  | Mean<br>(95% Confidence Interval)<sup>c</sup> |  |  |  |  |  |
| --- | --- | --- | --- | --- | --- | --- |
|  | Average Number of Individuals<sup>d</sup> |  |  | Monthly growth rate, % |  |  |
|  | Pre-Pandemic Level<sup>e</sup> | Pandemic Level<sup>e</sup> | Change in Level<sup>f</sup> | Pre-pandemic Trend<sup>g</sup> | Pandemic Trend<sup>g</sup> | Change in Trend<sup>h</sup> |
| All Payers | 212.28*<br>(196.85 to 227.71) | 210.72*<br>(199.53 to 221.91) | -1.56<br>(-18.95 to 15.83) | 4.83*<br>(3.40 to 6.26) | -0.70<br>(-1.84 to 0.44) | -5.53*<br>(-7.28 to -3.78) |
| Medicaid | 289.47*<br>(264.33 to 314.60) | 298.51*<br>(266.87 to 330.14) | 9.01<br>(-27.96 to 46.04) | 5.35*<br>(3.44 to 7.25) | -2.31<br>(-4.70 to 0.07) | -7.66*<br>(-10.42 to -4.90) |
| Commercial | 179.48*<br>(165.90 to 193.07) | 174.16*<br>(161.04 to 187.28) | -5.32<br>(-22.59 to 11.94) | 4.06*<br>(2.65 to 5.48) | 0.47<br>(-1.08 to 2.02) | -3.59*<br>(-5.53 to -1.66) |

<sup>a</sup>The dependent variable is the number of individuals filling prenatal buprenorphine/naloxone prescriptions, weighted by the number of pregnancies for a given month and payer. For example, the number of individuals filling prenatal buprenorphine/naloxone prescriptions on Medicaid in May 2019 is weighted by the number of pregnancies among patients on Medicaid in May 2019. This is to account for any seasonality in pregnancies or changing trends in pregnancies during the pandemic that may differ over time and by payer.
<sup>b</sup>The pre-pandemic period is defined as May 2019 through February 2020. The pandemic period is defined as April 2020 to December 2020.
<sup>c</sup>95% confidence intervals in parentheses below estimates.
<sup>d</sup>Represents the average number of individuals filling prenatal buprenorphine prescriptions per 100,000 pregnancies by payer.
<sup>e</sup>Predicted levels are calculated from a linear regression of monthly trends on the number of individuals filling prescriptions for buprenorphine/naloxone.
<sup>f</sup>The change in level is calculated as the coefficient on the term that measures the breakpoint in the interrupted time series (ITS) analysis.
<sup>g</sup>Predicted trends are calculated from a log-linear regression of monthly trends on the weighted number of individuals filling prescriptions for prenatal buprenorphine/naloxone.
<sup>h</sup>The change in trend is calculated as the coefficient on the interaction term in the interrupted time series (ITS) analysis where the dependent variable is log-transformed.
\* denotes $p \leq 0.05$

